# Disturbed laterality of non-rapid eye movement sleep oscillations in post-stroke human sleep: a pilot study

**DOI:** 10.1101/2023.05.01.23289359

**Authors:** Benjamin K. Simpson, Rohit Rangwani, Aamir Abbasi, Jeffrey M. Chung, Chrystal M. Reed, Tanuj Gulati

## Abstract

Sleep is known to promote recovery post-stroke. However, there is a paucity of data profiling sleep oscillations post-stroke in the human brain. Recent rodent work showed that resurgence of physiologic spindles coupled to sleep slow oscillations(SOs) and concomitant decrease in pathological delta(*δ*) waves is associated with sustained motor performance gains during stroke recovery. The goal of this study was to evaluate bilaterality of non-rapid eye movement (NREM) sleep-oscillations (namely SOs, *δ*-waves, spindles and their nesting) in post-stroke patients versus healthy control subjects. We analyzed NREM-marked electroencephalography (EEG) data in hospitalized stroke-patients (n=5) and healthy subjects (n=3) from an open-sourced dataset. We used a laterality index to evaluate symmetry of NREM oscillations across hemispheres. We found that stroke subjects had pronounced asymmetry in the oscillations, with a predominance of SOs, *δ*-waves, spindles and nested spindles in one hemisphere, when compared to the healthy subjects. Recent preclinical work classified SO-nested spindles as restorative post-stroke and *δ*-wave-nested spindles as pathological. We found that the ratio of SO-nested spindles laterality index to *δ*-wave-nested spindles laterality index was lower in stroke subjects. Using linear mixed models (which included random effects of concurrent pharmacologic drugs), we found large and medium effect size for *δ*-wave nested spindle and SO-nested spindle, respectively. Our results indicate considering laterality index of NREM oscillations might be a useful metric for assessing recovery post-stroke and that factoring in pharmacologic drugs may be important when targeting sleep modulation for neurorehabilitation post-stroke.

## Introduction

Stroke is a leading cause of motor disability world-wide, and despite advances in neurorehabilitation, there is a lack of widely adopted therapies that target plasticity and functional outcomes remain inconsistent ^1–3^. Sleep is known to play a major role in regulating plasticity^4–12^ and accordingly, there has been an interest in modulating sleep for stroke motor rehabilitation^13,14^. To optimize efforts to modulate sleep effectively, there is a need to better understand neural processing during sleep, as well as the effect of practical scenarios including patient co-morbidities and concurrent pharmaceuticals that may impact excitatory/inhibitory neural transmission. While a whole host of animal and human studies have shown that sleep can influence motor recovery post-stroke^2,14–23^, more work is needed to understand how sleep neurophysiology is affected in stroke, as well as inter-individual variations in human stroke patients. This has become all the more important with advances in our understanding of sleep neurophysiology linking nested non-rapid eye movement (NREM) oscillations to plasticity, motor memory consolidation, and motor recovery^4,6,14,24^.

Sleep-dependent neural processing is crucial for memory consolidation, which is the process of transferring newly learned information to stable long-term memory^9,25^. Initial investigations looked at sleep’s role in declarative memory^26,27^, but recent studies have underscored sleep’s role in motor skill consolidation^5,6,28^. Specifically, NREM sleep has been linked to reactivation of awake motor-practice activity and performance gains in a motor skill after sleep^4–6^. Increasingly, now there is consensus that this consolidation occurs during temporal coupling of sleep spindles (10– 16 Hz) to larger amplitude slow oscillations (SOs, 0.1–1Hz)^6,25,29–31^. Recent work in rodents has shown that these SOs nested with spindles decline immediately post-stroke and track motor recovery on a reaching motor task^14^. This work also showed that delta waves (*δ* waves, 1–4Hz), along with *δ* wave-nested with spindles increased post-stroke, and reduced during recovery.

These two nested oscillations (namely SO-nested spindles versus *δ* wave-nested spindles) had a competing role and pharmacological reduction of tonic γ-aminobutyric acid (GABA) neurotransmission shifted the balance towards restorative SO-nested spindles in the brain. The chief goal of our study was to see if NREM oscillations and their nesting were similarly affected post-stroke in human patients within a hospital setting, one within the framework of real-world stroke management, including co-morbidities and dissimilar pharmaceuticals that may modulate neural transmission. We also wanted to check for laterality of NREM oscillations in stroke and contralateral hemisphere and compare it to healthy subjects.

Our study showed that, acutely post-stroke, there is an increase in SOs and *δ* waves on stroke electrodes when compared to contralateral hemisphere electrodes, whereas healthy subjects had symmetrical density of these oscillations. We also found that spindles’ laterality was disturbed in stroke subjects with spindles being higher in the stroke hemisphere, except for one patient where spindles were higher in the contralesional hemisphere. This patient had a subcortical involvement in stroke. Our linear mixed effect model revealed that there was significant fixed effect of stroke vs contralateral electrodes for SOs and *δ* waves with overall medium effect sizes including random effects of concurrent pharmacologic drugs (propofol, dexamethasone, levetiracetam). We also observed a large effect size of linear mixed model for *δ* wave-nested spindles with random effects of pharmacologic drugs. Finally, we found that the ratio of SO-nested spindles laterality index to *δ*-wave-nested spindles laterality index was lower in stroke subjects when compared to healthy subjects. Together, our work suggests that laterality of NREM sleep oscillations could be a useful marker for physiological sleep activity post-stroke, and that acute stroke care management should incorporate pharmacologic drug interactions and their effects on laterality of ‘restorative’ sleep oscillations. This may help inform a personalized approach to sleep modulation for neurorehabilitation.

## Patients and Methods

### Ethics, consent and permissions

This research was conducted in accordance with and approval of the Cedars-Sinai Medical Center Institutional Review Board (IRB). All research participants and/or their surrogates provided informed consent to participate in the study.

### Inclusion/exclusion criteria

Retrospective chart review of the Cedars Sinai EEG database was done to identify patients with acute middle cerebral artery strokes (MCA strokes; with high probability of stroke lesion affecting sensorimotor regions in the brain) who also received EEG monitoring as part of their hospital stay. We selected patients who received EEG in the acute period (2-3 days) post-stroke. Other inclusion criteria were that this should be the first stroke for the patient, they should be within 50-80 years of age, and the patients should not have any sleep disorders or circadian /diurnal rhythm disruption. Subjects were excluded if they met the following criteria: currently pregnant or diagnosed with uncontrolled medical conditions. Five patients were retrospectively identified for this study, with notable limited availability of EEG studies done within 2-3 days after an MCA distribution stroke. Of the 5 patients, 3 were female and 2 male, all within the age range of 50-80 years old (see **Table 1** for other details regarding demographic and clinical information). Indications for EEG were universal for altered mental status after acute stroke. Unlike all other patients, P4 had subcortical involvement in stroke. It is important to note that spindle oscillations are postulated to have have a subcortical (thalamocortical) origin^32^. Also of note is that P5 had a hemorrhagic stroke (ruptured right MCA aneurysmal stroke) and was on norepinephrine due to shock which improved within 24 hours. P2 had partial status epilepticus involving the right temporal lobe. We excluded seizure related epochs based on manual inspection of recordings. This inspection was done by epileptologist (CMR) and seizures were excluded based on no evolving seizure pattern across electrodes (10-20 EEG system). Hence, all of our presented data was from sleep periods in all the five patients (even in the patient with status epilepticus). An average of ∼5.9801 ± 1.2563 hours (or 358.8036 ± 75.4 mins, mean ± s.e.m.) of NREM sleep was identified and analyzed in each of the five patients. Additionally, we analyzed a dataset of 3 healthy subjects from Cox et. al, Sleep medicine reviews, 2020^33,34^ where average NREM sleep analyzed was 3.0652 ± 0.1396 hours (or 183.9100 ± 8.3773 mins) for 3 subjects. We were not able to analyze REM/ wake periods in these recordings due to inability to differentiate these periods due to the lack of EMGs/ video recordings.

**Table 1.** Patient clinical information. Top to bottom, information for five patients P1 to P5. Patient age, sex, race/ethnicity, stroke location, NIHSS, days from stroke when the EEG data was acquired, associated co-morbidities, sleep disorders, circadian rhythm disruption, alcohol and smoking substance consumption status, and concurrent medications during EEG recording are specified. Abbreviations; NIHSS: National Institutes of Health Stroke Scale; R/ L MCA: Right/ left middle cerebral artery; COVID: Coronavirus disease - 2019; ESRD: End-stage renal disease; HFrEF: Heart failure with reduced ejection fraction; HypoT: hypothyroidism; ASA: Acetylsalicylic Acid (Aspirin); N/A: not available. Patient groups: **blue**: patients in propofol medication group (Group-1); **orange**: patients in levetiracetam medication group (Group-2); **green**: patients in other medication group (Group-3).

| Patient | P1 | P2 | P3 | P4 | P5 |
| --- | --- | --- | --- | --- | --- |
| Age | 56 | 68 | 51 | 56 | 52 |
| Sex | F | F | M | M | M |
| Race/ethnicity | Hispanic | White/Caucasian | Hispanic | Black/African-american | White/Caucasian |
| Stroke location | R MCA | R MCA | L MCA | R MCA | R MCA |
| NIHSS | 3 | N/A | 21 | N/A | N/A |
| Time of recording after stroke | 2 days | 2 days | 3 days | 3 days | 3 days |
| Comorbidities | COVID | Partial status epilepticus (right temporal) | ESRD, HFrEF | Pituitary macroadenoma, Central hypoT | Ruptured R MCA aneurysm |
| Sleep disorders (e.g. obstructive sleep apnoea) | No | No | No | No | No |
| Circadian rhythm disruption | No | No | No | No | No |
| Alcohol | Yes | No | N/A | No | No |
| Smoking | No | No | N/A | No | No |
| Rx (concurrent) | Propofol gtt<br>Dexamethasone<br>Remdesivir | Levetiracetam<br>Acyclovir<br>Vancomycin<br>Cefepime | ASA/Plavix | ASA<br>Levothyroxine | Levetiracetam<br>Levophed |

### EEG analysis and identification of NREM oscillations

Patients with overnight EEG studies 2 to 3 days post-stroke, with appropriate clinic follow up, were included. The data, obtained by a Natus Xltek EEG and Sleep System, was de-identified and made compatible for analysis with MATLAB. Each 30-second epoch was individually and manually marked for NREM sleep by an expert scorer (C.M.R. and B.K.S.). EEG epochs were analyzed for NREM sleep in a bipolar montage. The following analysis was done with EEG data in a referential montage, referenced to the auricle electrodes. Spindles, SOs, and *δ* waves were extracted from these NREM epochs using custom code in MATLAB (details below). This allowed for the identification of specific sleep waveforms and how they nested temporally and topographically during NREM sleep. An emphasis was placed on assessing spindles and their nesting to SOs and *δ* waves. Topographical maps of the average density of these sleep waveforms allowed us to visualize the average densities with respect to electrode location, especially their lateral symmetry between hemispheres.

From the healthy subjects dataset, we used the *common linked mastoids* referenced data^33^ and analyzed NREM sleep. We also identified and only selected 20 electrode channels in similar locations as stroke patient data for further analysis (because the healthy subject data had more electrodes than our stroke patient dataset). Similar to stroke EEG data, spindles, SOs, and *δ* waves were extracted from these NREM epochs using custom code in MATLAB and analyzed.

#### EEG Data processing

For stroke patients, NREM-marked EEG data from all channels was referenced with respect to the average of the auricular electrodes (A1 & A2, **Fig. 1A**) while the heathy control dataset had common linked mastoids referenced EEG data. Any high amplitude artifact in the differential EEG signal was removed. This data was filtered into the frequency ranges of 0.1-4 Hz for *δ*/SOs identification and 10-16 Hz for spindle detection. We used previously used methods for automatic detection of these NREM oscillations^6,14,35^. For ***δ/SOs detection***, signal was first passed through a 0.1 Hz high-pass filter and then a 4 Hz low-pass Butterworth filter. All positive-to-negative zero crossings, previous peaks, following troughs, and negative-to-positive zero crossings were identified. A wave was considered a *δ* wave if its trough was lower than the negative threshold and preceded by a peak that was lower than the positive threshold, within 500 ms (**Fig. 1B, E, H**). SOs were classified as waves with troughs lower than a negative threshold (the bottom 40 percentile of the troughs) and preceding peaks higher than a positive threshold (the top 15 percentile of the peaks; **Fig. 1C, F, I**). Duration between peaks and troughs was between 150 ms and 500 ms. For ***spindle detection***, EEG data was filtered using a 10 Hz high-pass Butterworth filter and a 16 Hz low-pass Butterworth filter. A smoothed envelope of this signal was calculated using the magnitude of the Hilbert transforms with convolving by a Gaussian window (200 ms). Epochs with signal amplitude higher than the upper threshold (mean, µ + 2.5*s.d., σ) for at least one sample and amplitude higher than the lower threshold (µ + 1.5*σ) for at least 500 ms were considered spindles (**Fig. 1 D, G, J**). The lower threshold was used to define the duration of the spindle. Nested SO-spindles (similar to *k*-complexes studied in humans) were identified as spindle peaks following SO peaks within 1.5 s duration (**Fig. 1K**). The same criterion was used to identify *δ* wave-nested spindles (**Fig. 1L)**.

**Figure 1.**
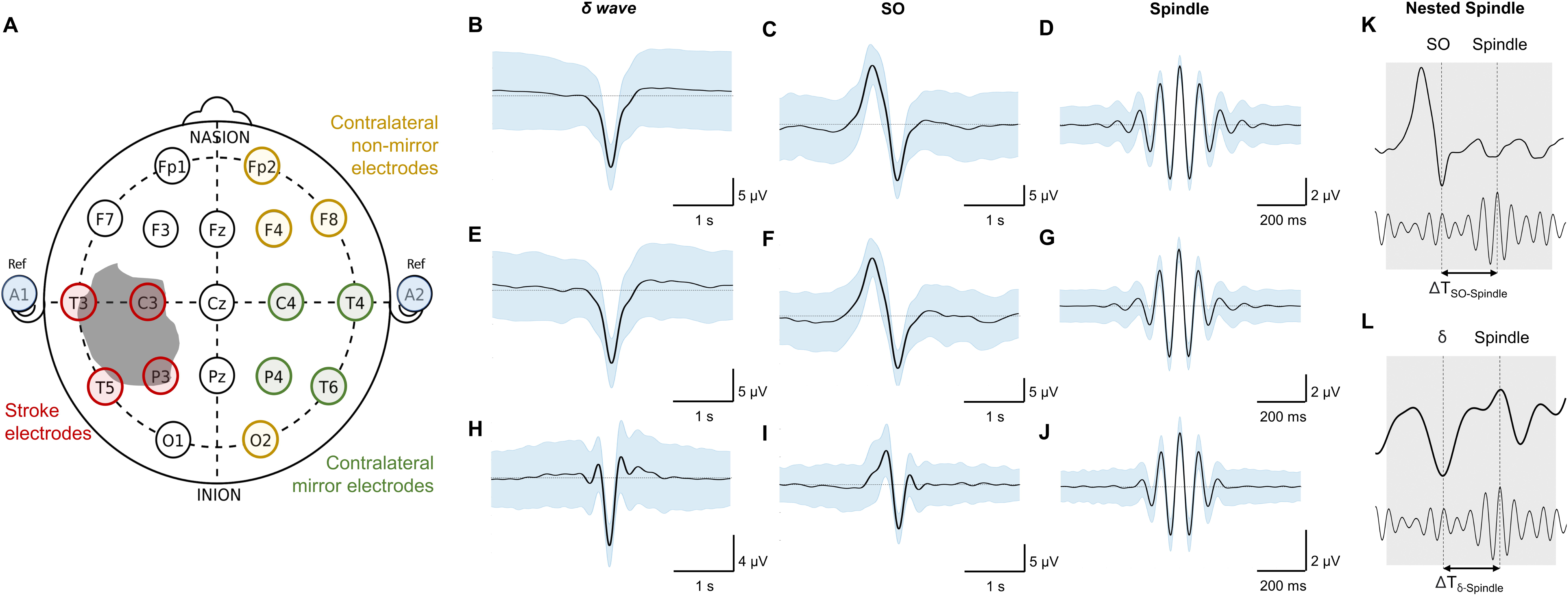
Stroke versus contralateral mirror/non-mirror electrode assignment and NREM sleep oscillations. ***A*,** 10–20 system for EEG (used in stroke patients) showing locations of all electrode locations recorded with an illustration of stroke. Grey shaded area shows a representative stroke perilesional region. Blue shaded circles represent auricular electrodes (A1, A2) that were used for referencing in the stroke patients. Red circles indicate identified *stroke electrodes* based on proximity to the perilesional area. Green circles indicate identified *contralateral mirror* (CM) *electrodes* which are contralateral and mirrored to identified *stroke electrodes. Yellow* circles indicate identified *contralateral non-mirror* (CNM) *electrodes* which are electrodes other than *contralateral mirror (CM) electrodes* in non-stroke hemisphere*. **B***, Mean *δ–* wave along with s.e.m. (standard error of mean) bands (blue) for all identified *δ–*waves from an example *stroke electrode* channel from EEG data recording for one stroke patient. ***C*,** Same as ***B*** for SO waveforms. ***D,*** Same as ***B*** for spindle waveforms. ***E*, *F, G,*** Same as ***B, C, D*** for one example *contralateral mirror* electrode channel for a stroke patient. ***H, I*, *J*** Same as ***B, C, D*** for one example channel for a healthy subject. All waveforms are centered around the detected states. ***K*,** Illustration of SO-spindle nesting. Nesting window was –0.5 to +1.0 s from SO’s UP state as shown. ***L*,** Illustration of *δ–*wave-spindle nesting. Nesting window was –0.5 to +1.0 s from δ UP state as depicted.

##### Data Analysis

We generated topographical maps of these different waveforms using *plot_topography* function in MATLAB^36^ as shown in **Fig. 2**. The patients were separated into 3 different groups based on concurrent medications, as detailed in **Table 1**. Patient 1 was assigned to Group 1, who was on continuous propofol and dexamethasone injections every four hours. Group 2, patients 2 and 5, was administered levetiracetam (Keppra) twice daily; and Group 3, comprised of patients 3 and 4, was not on medications known to significantly modulate excitatory/inhibitory neural transmission.

**Figure 2.**
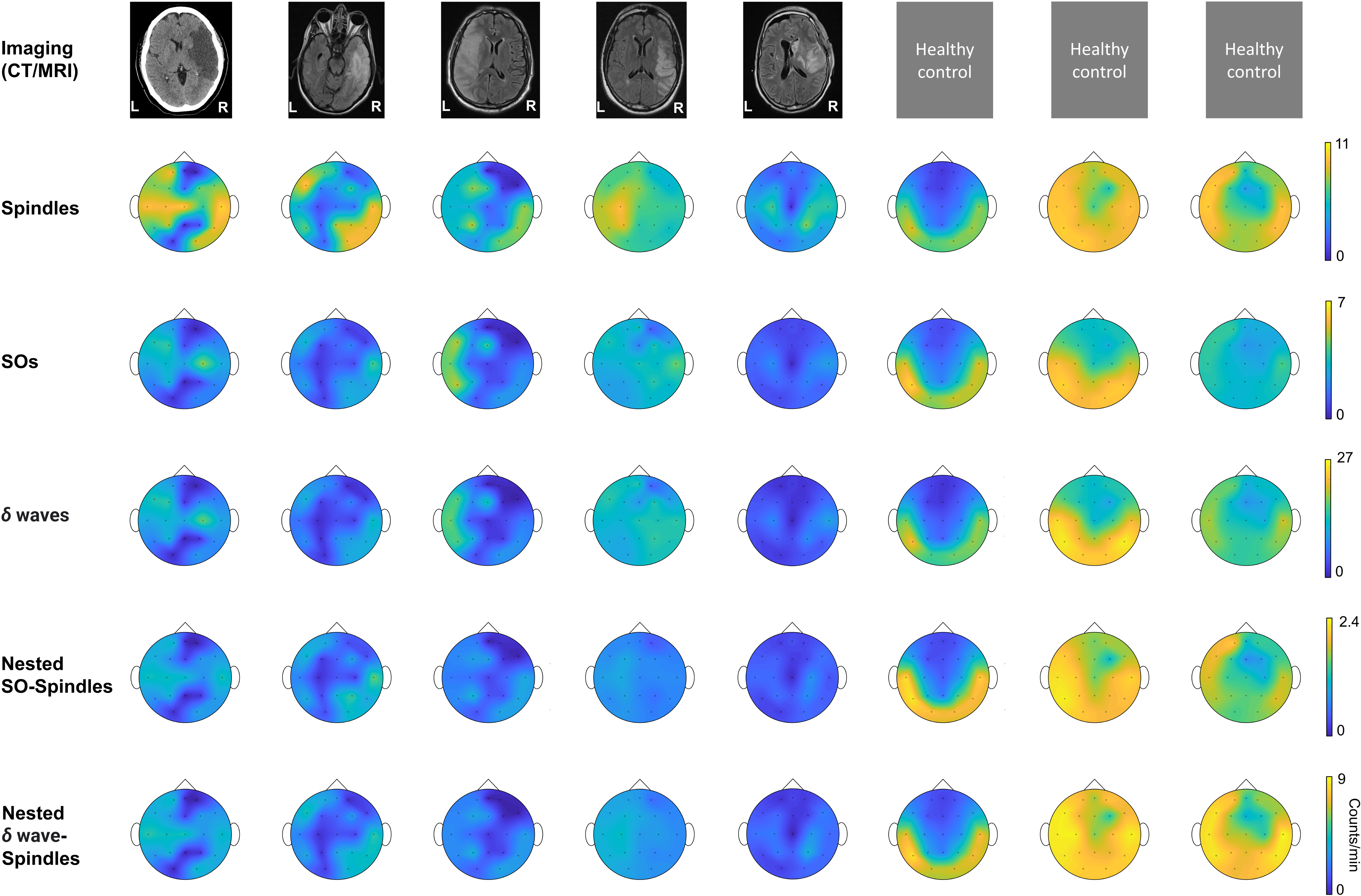
Imaging data and topographical density plots for different NREM oscillations. Top to bottom: ***Imaging data***: CT (computed tomography) image for patient P1, T2 sequences of MRI (magnetic resonance imaging) images for patients P2 to P5; no imaging data available for healthy subjects (P6 to P8). Radiologic imaging has been flipped horizontally to align with topographic density maps; *i.e.*, image left, and right are ipsilateral to patient left and right. Left and right are marked in imaging figures (P1-P5) and apply to density topographical maps below them; ***Topographical maps*** for detected ***spindle*** density (count/min) during NREM sleep for all subjects; ***Topographical maps*** for detected ***SO*** density (count/min) during NREM sleep for all subjects; ***Topographical maps*** for detected ***δ waves’*** density (count/min) during NREM sleep for all subjects; ***Topographical maps*** for detected ***nested SO-spindle*** density (count/min) during NREM sleep for all subjects; ***Topographical maps*** for detected ***δ wave-nested***-***spindle*** density (count/min) during NREM sleep for all subjects. Color map shown at right for all the panels in a row.

Perilesional electrodes were identified by analyzing post-stroke MRI and CT brain imaging. We marked ‘Stroke electrodes’ as the electrodes covering the perilesional region of the brain as shown in **Fig. 1A**. The mirror opposite electrodes on the contralateral side were marked as “Contralateral mirror (CM) electrodes’ for further analysis (**Fig. 1A**). The non-mirror opposite electrodes on the contralateral side were marked as “Contralateral non-mirror (CNM) electrodes’.

We compared the symmetry in NREM oscillations’ density across hemispheres for stroke patients and healthy control using a laterality index (**Fig. 3A-F**). Laterality index of 1 meant the average density being analyzed for electrode locations selected across hemisphere is exactly same. For stroke patients, laterality index is defined as the ratio of mean of stroke electrodes’ NREM densities to all contralateral electrodes’ NREM densities. For healthy subjects, laterality index is defined as the ratio of mean of left hemisphere electrodes’ NREM densities to right hemisphere electrodes’ NREM densities. We also compared the ratio of SO-nested spindles laterality index to *δ* wave-nested spindles laterality index for stroke vs healthy subjects.

**Figure 3.**
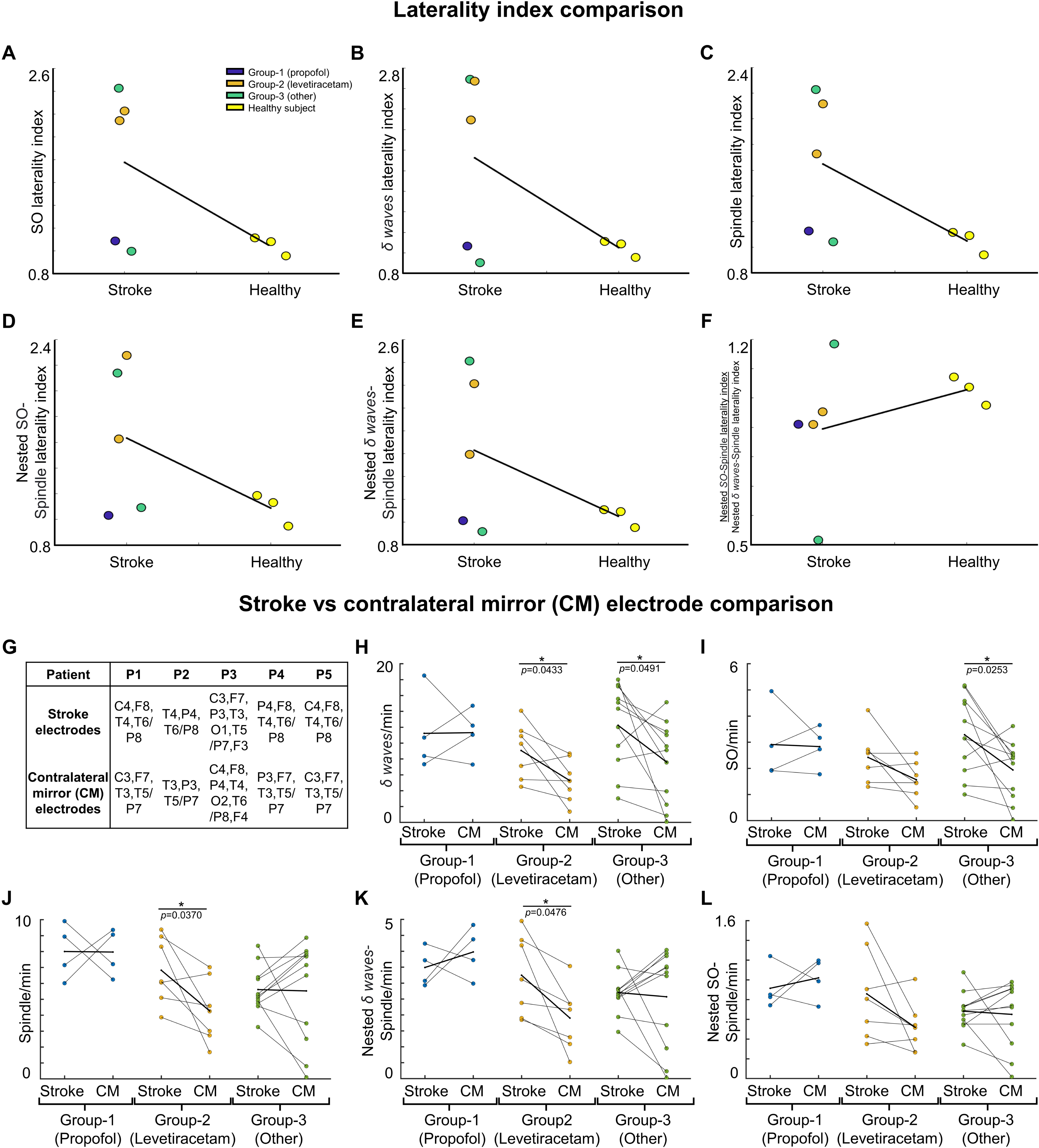
NREM oscillations’ laterality in stroke patient’s vs healthy controls; and NREM oscillations’ densities for different patient groups on stroke verses contralateral mirror (CM) electrodes. For stroke patients’ laterality index (LI) is defined as ratio of mean of stroke electrode NREM densities to all contralateral electrode NREM densities. For healthy subjects’ laterality index is defined as ratio of mean of left hemisphere electrode NREM densities to right hemisphere electrode NREM densities. *A,* LI for SO density for stroke patients and healthy controls. Black line connects the mean of stroke and control group. Dots represent different patients/subjects; **blue dots**: Patients in propofol medication group; **orange dots**: Patients in levetiracetam medication group; **green dots**: Stroke patients in other medication group; **yellow dots**: Healthy subjects. *B,* Same as *A* for *δ* wave density LI. *C,* Same as *A* for spindle density LI. *D,* Same as *A* for nested SO-spindle density LI. *E,* Same as *A* for Nested *δ* wave-spindle density LI. *F,* Ratio of LI for nested SO-spindle density and nested *δ* wave-spindle density. *G*, Table showing selected *stroke* and *contralateral mirror electrodes* (CM) for all patients. *H*, Comparison of *δ wave* density (count/min) on *stroke versus CM electrode*s for patients on different medications. Thick black line shows the mean values within the group. Thinner black lines join pair of stroke and CM electrode. Dots represent the NREM oscillations’ density for single electrode. *I*, Same as *H* for SO density. *J*, Same as *H* for spindle density. *K*, Same as *H* for nested *δ* wave-nested spindle density. *L*, Same as *H* for SO-nested spindle density. *: statistically significant *p* values for two-tailed *t*-test.

### Statistical Analysis

We performed a linear mixed effect analysis for all patients comparing the *Stroke electrodes* density vs Co*ntralateral (CM/CNM) electrodes* density for different waveforms using the *fitlmematrix* function in MATLAB. The linear mixed effect model was fitted by maximum likelihood using the formula below **(1)** for all the different waveforms identified during EEG data processing. Medication groups were defined as the 3 groups as mentioned earlier. This model considers fixed effects of stroke vs contralateral (CM/CNM) electrodes, and the random effect of electrodes and medication groups depending on the patient and can be represented as following equation:

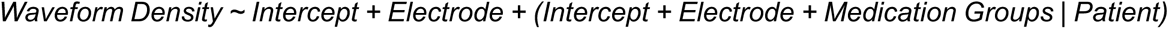

The above formula/equation is written in a format similar to documentation for *fitlmematrix* Matlab function. We compared the *Stroke electrodes* density vs *contralateral (CM/CNM) electrodes* density within each medication group using a two-tailed *t*-test. Contralateral electrodes chosen were the exactly mirrored electrodes (**Fig. 3G–L**) or non-mirrored (**Supp. Fig. 2A-F**). One-way ANOVA was used to compare the stroke electrodes’ NREM oscillations’ density of the 3 different medication groups.

We calculated r-squared (*R*^2^) and the Cohen’s *d* values for the overall linear mixed effect model generated. However, the p-values were specifically assessed for fixed effect of electrodes (stroke vs CM/CNM). Cohen’s *d* was used to evaluate if the nested data (all data combined) for NREM oscillations had a small, medium or large experimental effect (Cohen’s *d* = 0.20, 0.50 or 0.80, respectively)^37^. Effect size indicates if such research findings have practical significance. Metrics such as Cohen’s *d* are better at the planning stage for pilot studies such as the one here to determine optimal sample sizes for sufficient power in bigger clinical trials^38^. We have summarized the linear mixed effects models results in a table in the Supplementary Information.

## Results

One of the limitations of retrospectively analyzing EEG data gathered from clinical EEG was the heterogeneity encountered across the subjects studied, a contrast from the controlled setting of related rodent studies. Knowing this, we found that one important similarity across the study population was the indication for EEG: concern for underlying seizure in the setting of altered mental status and recent hemispheric stroke. Accordingly, the patients were all hospitalized and our analysis benefited from close pharmacologic documentation. We observed stark differences in laterality of NREM oscillations in stroke patients. We observed higher SOs, δ waves, spindles and spindles nested to SOs and δ waves in the stroke hemisphere. For the patient with subcortical involvement in stroke, we observed decrease in spindles in the stroke hemisphere. We also observed random effects of concurrent medications, particularly medications that influenced neural transmission. P1 was noted to be on continuous infusion of propofol (<10 mcg total) and infusions of dexamethasone every 4 hours. P2 and P5 were treated with levetiracetam 500mg twice daily. P2 was also on acyclovir which was discontinued after cerebrospinal fluid (CSF) tested negative for meningitis; and P5 was administered nonepinephrine due to being in shock acutely and improved within 24 hours. P3 and P4 were not given propofol, dexamethasone, or levetiracetam.

### NREM oscillation densities symmetry is disturbed acutely in stroke

We found that stroke patients had laterality differences (densities were higher or lower in stroke hemisphere) for all NREM oscillations, while the healthy subject NREM oscillation density looked more symmetrical across hemispheres (**Fig. 2**). Comparing the laterality index (LI) (as defined in methods), we found that the LI was closer to 1 on average for healthy subjects and had low variance but there were differences in stroke patients. SO density LI, stroke: 1.7761 ± 0.3387 and healthy: 1.0520 ± 0.0587 (**Fig. 3A**). For *δ wave* density LI, stroke: 1.9251 ± 0.4368 and healthy: 1.0521 ± 0.0592 (**Fig. 3B**). For spindle density LI, stroke: 1.6478 ± 0.2736 and healthy: 1.0498 ± 0. 0.0669 (**Fig. 3C**). For SO-nested with spindles LI, stroke: 1.6325 ± 0.2876 and healthy: 1.0884 ± 0.0882 (**Fig. 3D**). For *δ* wave-nested with spindles LI, stroke: 1.6299 ± 0.3387 and healthy: 1.0525 ± 0.0608 (**Fig. 3E**). The ratio of nested SO-spindles LI and *δ* wave-nested spindle LI, stroke: 0.8948 ± 0.1201 and healthy: 1.0280 ± 0.0344 (**Fig. 3F**).

### SO and *δ* wave density increased in perilesional electrodes

Next, we wanted to closely look at stroke-affected electrodes in stroke patients vis-à-vis the contralateral hemisphere electrodes in same stroke subjects. In the contralateral hemisphere, we looked at exactly mirrored electrodes (CM, as defined in the methods; **Fig. 3G**), or non-mirrored electrodes (CNM, as previously in methods; **Supp. Fig. 2A**). Consistent with previous reports, we found that stroke electrodes had higher < 4 Hz oscillations (**Fig. 3H,I**; and **Supp. Fig. 2B,C**)^39^. Our mixed-effects model showed a significant fixed effect of stroke vs CM and CNM electrodes for a subset of NREM oscillations and overall medium to large effect sizes which included random effects of concurrent pharmaceuticals. Overall, we observed higher *δ* wave density in the perilesional electrodes (**Fig. 3H; Supp. Fig. 2B; Supp. Table 1** and **2** provide statistical details for stroke versus CM or CNM: p-value is provided for the fixed effect (’electrode’), *R*^2^ and Cohen’s *d* are for the overall model with fixed and random effects, coventions same henceforth). Our comparison of laterality index of SOs and *δ* wave showed that LI was higher in stroke patients compared to healthy subjects: Mean LI SOs, stroke: 1.7761 ± 0.3387 and healthy: 1.0520 ± 0.0587; Mean LI *δ* wave, stroke: 1.9251 ± 0.4368 and healthy: 1.0521 ± 0.0592. We also observed that the stroke patients on propofol and dexamethasone (Group-1) and Group-3 both had higher *δ* wave density on stroke electrodes than the levetiracetam group (**Fig. 3H** and **Supp. Fig. 2B**; stroke electrodes’ *δ* wave density-Group 1: 11.2315 ± 2.5270 counts min^−1^ (mean ± s.e.m.); Group 2: 9.0653 ± 1.3206 counts min^−1^; Group 3: 12.2454 ± 1.5866 counts min^−1^, see **Supp. Table 3** for details). Levetiracetam (Group-2) and Group-3 showed high density of *δ* waves in the stroke electrodes vs CM/ CNM electrodes (**Fig. 3H** and **Supp. Fig. 2B**). For SOs as well, there was a significant fixed effect of stroke vs contralateral electrodes (**Fig. 3I; Supp. Fig. 2C**; **Supp. Table 1** and **2** provide p-values and Cohen’s d). When we looked at stroke patients with different drugs, we observed that the patients in Group-1 did not show a significant difference between stroke or contralateral electrode SO density; patients in Group-2 showed elevation in SO on stroke electrodes when compared to CM electrodes (**Fig. 3I**). The patients in Group-3 showed more SOs on stroke electrodes when compared for CM/CNM electrodes (**Fig. 3I; Supp. Fig. 2C**; stroke electrodes’ SO density: Group 1: 2.9134 ± 0.7147 counts min^−1^; Group 2: 2.4204 ± 0.3701 counts min^−1^; Group 3: 3.2858 ± 0.4464 counts min^−1^; see **Supp. Table 3** for details).

For spindle oscillations, LI was higher in stroke patients (Mean LI spindles, stroke: 1.6478 ± 0.2736 and healthy: 1.0498 ± 0. 0.0669). Interestingly, in one patient with subcortical involvement with stroke (P4), spindles were higher in contralesional hemisphere (**Fig. 3J**). Linear mixed-effects model did not show a significant fixed effect of spindle density on stroke versus contralateral electrodes, however, overall it was a medium effect based on the Cohen’s *d* (**Fig. 3J** and **Supp. Fig. 2D;** see **Supp. Table 1** and **2** for p-value and Cohen’s *d*). Upon looking at different stroke patients, spindle density was the highest on the stroke electrodes in the patient in Group-1 (8.0037 ± 0.8763 counts min^−1^), followed by the patients in Group-2 (6.8267± 0.7872 counts min^−1^), and then patients in Group-3 (5.6120 ± 0.4366 counts min^−1^) (**Fig. 3J** and **Supp Fig. 2D**; see **Supp. Table 3** for details).

### *δ* wave-nested spindles and SO-nested spindles

Next we wanted to look at nested oscillations, namely, δ wave-nested spindles and SO-nested spindles oscillations that were shown to have a competing role recently and inverse trend during stroke recovery^6,14^. LI for both nested oscillations was higher in stroke subjects: Mean LI SO-nested spindle, stroke: 1.6325 ± 0.2876 and healthy: 1.0884 ± 0.0882; Mean LI *δ* wave-nested spindle, stroke: 1.6299 ± 0.3387 and healthy: 1.0525 ± 0.0608. While the mixed effects models on *δ* wave-nested spindles and SO-nested spindles did not show a significant difference between stroke and contralateral electrodes, overall these models still had large and medium effect sizes, respectively (**Supp. Table 1** and **2**, **Fig. 3K** and **Supp. Fig. 2E,** *δ* wave-nested spindle density on stroke electrodes: Group-1: 3.4911 ± 0.3032 counts min^−1^; Group-2: 3.2476 ± 0.4770 counts min^−1^; Group-3: 2.7031 ± 0.2032 counts min^−1^, also see **Supp. Table 3**; *SO*-nested spindle density on stroke electrodes: Group 1: 0.9150 ± 0.1111 counts min^−1^; Group 2: 0.8583 ± 0.1751 counts min^−1^; Group 3: 0.6828 ± 0.0576 counts min^−1^; see **Fig. 3L**; **Supp. Fig. 2F**; and **Supp. Table 3**). Notably, the ratio of SO-nested spindle LI to *δ* wave-nested spindle LI was lower in stroke subjects compared to heathy subjects (Mean LI ratio, stroke: 0.8948 ± 0.1201 and healthy: 1.0280 ± 0.0344), which indicates relatively increased *δ* wave-nested spindles when compared to SO-nested spindles (the oscillations that have a competing role in forgetting vs strengthening, respectively) in the perilesional areas for stroke brain compared to healthy brain.

Together, these results show that lateral symmetry of NREM oscillations is disturbed in stroke (**Fig. 3A-F**), when compared to healthy subjects. These results also indicate that there is an elevation of SO, *δ* wave, spindles, and spindle nesting to SOs or *δ* waves in the perilesional areas post-stroke.

## Discussion

Our results show that, post-stroke there is a disturbance in laterality of NREM sleep oscillations between ipsilesional and contralesional hemisphere. Interestingly, hemispherical differences in these nested oscillations were less pronounced in healthy subjects, and oscillations appeared mostly symmetric. We used a laterality index for comparing NREM oscillations, especially nested oscillations, *i.e., SO*-nested spindle oscillations and *δ* wave-nested spindle oscillations. These two nested oscillations were recently demonstrated to have a competing role during stroke recovery in preclinical setting. Our results here might help improve neuromodulation of sleep for rehabiliation that targets improvement in laterality index of NREM sleep oscillations. While our findings are preliminary in a small pilot dataset, these findings report an interesting effect size, suggesting a roadmap for delineating pathological sleep in larger cohorts and optimal therapeutic modulation to promote recovery.

### Sleep and plasticity post-stroke

Preclinical and clinical studies that have evaluated local-field potentials (LFPs) in animals^40,41^ and EEG in human patients^22,42,43^ have found increased low-frequency power during awake, spontaneous periods after a stroke. These studies postulate that this increased low-frequency activity could be a marker of cortical injury and loss of subcortical inputs^44^. Our findings on increased SOs and *δ* waves on stroke electrodes are indicative of similar phenomena. We also found an increase in SO-nested spindles and *δ* wave-nested spindles on stroke electrodes along with a lower ratio of SO-nested spindle LI/ *δ* wave-nested spindle LI (**Fig. 3F**). There is growing evidence that temporal coupling of spindles to SOs is a primary driver of sleep-related plasticity and memory consolidation^6,30,31,45–48^. SO-nested spindles are linked to spike-time dependent plasticity^49^. These events are also related to reactivation of awake experiences^30,47,50^. Importantly, disruption of this coupling can impair sleep-related memory consolidation of awake experiences^6^. This same work showed that SO-nested spindles and *δ* wave-nested spindles compete with each other to either strengthen or forget a memory. Our results indicate that balance of SO-nested spindle density and *δ* wave-nested spindle density is disturbed hemispherically in stroke patients compared to healthy subjects. These might be related to impaired sleep-processing that may impact recovery. Interestingly, we observed large-to medium effect sizes in our mixed-effects models for *δ* wave-nested spindle and SO-nested spindle where we considered fixed effects of electrodes and random effects of drugs and patients. It is worth noting that drugs like propofol can impact such nested sleep oscillations^51,52^. It may be important to consider effects of drugs on sleep oscillations when modulating sleep for stroke recovery.

### Propofol and Levetiracetam: effect on sleep

We made some observations on different medications that stroke patients received during sleep EEG recordings. Group-1 patient received propofol, which is one of the most commonly used anesthetics in neurologic intensive care units after stroke or traumatic brain injury^53^. It exerts its action by potentiating the activity of chloride currents through GABA receptors while blocking voltage-gated sodium channels^54–56^. The patient on propofol received less than 10 mcg dose of propofol at which does it is not known to impact sleep^57,58^. Group-2 patients received levetiracetam (keppra), which is a relatively newer anti-seizure drug. The exact mechanism for its anti-seizure mechanism is unclear, but it is believed to exert its effect through synaptic vesicle glycoprotein 2A^59^. Through this mechanism, levetiracetam is capable of modulating neurotransmission by inhibiting calcium currents^60^. A study has shown that levetiracetam has minimal effect on sleep parameters like total sleep duration, sleep latency, and sleep efficiency in both healthy humans and partial epilepsy patients^61^. However, observations have been made that levetiracetam can reduce motor activity and cause daytime drowsiness in patients^61,62^. Propofol, by its GABAergic action, causes greater loss of faster frequencies during induction with a shift in alpha frequencies to the frontal regions that reverses post-awakening^63–65^. Since our mixed-effects model found large to medium effects when considering random effects of drugs on all NREM oscillation, it may be useful to explore effects of drugs on NREM sleep densities with larger patient cohorts in the future. Since our data shows disturbed laterality of NREM oscillations, different drugs that stroke patients are on might need to be factored in, when restoring physiological sleep oscillations post-stroke.

### Sleep processing and stroke rehabilitation

Recent rodent work profiled SO-nested and *δ* wave-nested spindles during the course of stroke recovery and found links between these nested structures and motor performance gains during recovery^6^. This work specifically looked into gains on reach-training, but clinical rehabilitation approaches can be varied^66–68^. It is likely that the sleep features of nested oscillations and their putative pathological or physiological roles need to be factored in when considering timing for rehabilitation, irrespective of training type. Previous human and rodent studies have also suggested critical periods in training can offer long-term benefits^69–71^. Past studies that have found awake low-frequency power in stroke patients might be related to our findings of increased SO and *δ* waves densities. Future studies where EEG data is captured over the long term may delineate a transition of *δ* wave LI, SOs LI, *δ* wave-nested spindles LI (pathological sleep) and SO-nested spindle LI (physiological sleep), and its relation to critical periods post-stroke for optimal timing of rehabilitation. For example, SO-nested spindles LI and *δ* wave-nested spindles LI proportions between hemispheres could be targeted to be brought closer to unity as in healthy subjects, to accelerate recovery.

### Modulation of sleep as a therapeutic intervention

The results we have presented can form the basis of translational studies in the future that target modulation of sleep post-stroke. Animal studies have suggested that modulation of GABAergic transmission (specifically GABA_A_-receptor mediated tonic inhibition) in the perilesional cortex can serve as a therapeutic target to promote recovery, and that blocking of GABA_A_-mediated tonic inhibition promoted motor recover maximally in the first 1 to 2 weeks post-stroke^72,73^. Both short-term (acute) and long-term chronic infusion of GABA_A_ inhibiting compounds have been tested, and long-term infusion was shown to be better^72^. Long-term pharmacologic modulation as shown by Clarkson and colleagues may be essential to achieve observable motor benefits in human patients. Benefits of long-term infusion include effect of the drug not only with rehabilitation-specific online (awake) training, but also during offline memory consolidation during sleep.

Studies such as ours can also help guide electric stimulation-based neuromodulation for augmenting recovery. SOs and *δ* waves can be easily monitored using EEG in stroke patients. Non-invasive brain stimulation during sleep^30,47,74,75^ can be used to modulate specific NREM oscillations. Invasive stimulation approaches such as epidural stimulation^76^ can also focus on sleep state to optimize sleep neural processing. Similar approaches have shown that direct epidural motor cortical electric stimulation can enhance awake performance and neural activity^77,78^ and epidural stimulation of subcortical regions can also modulate low-frequency oscillations in the motor cortex^79^, however such approaches have not been applied during sleep. A recent study suggested modulating UP states during sleep can enhance recovery^18^. It is plausible that future approaches targeting sleep, when delivered in a closed-loop fashion, optimize both awake task performance and its consequent sleep processing, and may lead to greater long-term benefits during rehabilitation. Indices such as laterality index that we pursued here may serve a utilitarian purpose in long-term sleep evaluation post-stroke with different treatments. Our pilot observations here also suggest that concurrent pharmacologic drugs may affect NREM oscillations and hence they should also be considered when personalizing sleep stimulation.

### Limitations

One of the limitations of our study is the lack of a link between sleep architecture and motor status. Future work that studies sleep over several days post-stroke and assesses motor functionality longitudinally may find more robust links between sleep processing and related gains in motor performance. It is also possible that, with more effective task performance and associated awake neural dynamics^77,78,80^, efficacy of sleep may change. Precise disruption of sleep processing, specifically SO-spindle coupling in healthy animals, was sufficient to prevent offline performance gains, even when awake task learning was robust^6^. This work also showed that precise modulation of the extent of sleep spindle-SO coupling in healthy animals could either enhance or impede sleep processing. While extension of this work in stroke animals has shown SO-spindle nesting resurges with recovery^14^, future animal studies that modulate sleep microarchitecture can study if artificial manipulation of SO-nested spindles or *δ* wave-nested spindles after stroke are sufficient to enhance or impair motor recovery. Our work here showed that both SO-nested spindles and *δ* wave-nested spindles increased in stroke affected hemisphere acutely post-stroke. Future work that monitors these oscillations for longer periods will be needed to assess if SO-nested spindles should increase relatively to *δ* wave-nested spindles for better recovery in human stroke patients.

One more limitation of this study is the limited sample size with varying lesion location and size. But ours is a pilot study. While we focused on getting patients with cortical lesions, sleep may have been impacted differently for one patient with a primarily subcortical stroke. For example, a stroke in the white matter that impacts thalamocortical networks may impact spindles (as we saw in our P4, with higher spindles in contralesional hemisphere), believed to have a thalamocortical origin. Future work with larger sample sizes and incorporation of motor task rehabilitation training and drug manipulation, may provide stronger links to engineer sleep to benefit motor recovery post-stroke.

## Data Availability

All data produced in the present study are available upon reasonable request to the authors

## Author Contributions

B.K.S., R.R., C.M.R. and T.G. contributed to the design of the study. R.R., B.K.S. and A.A. contributed to the analysis of the data. B.K.S., J.M.C. and C.M.R., contributed to the acquisition of data. B.K.S., R.R., C.M.R. and T.G. contributed to the interpretation of the data. B.K.S., R.R., C.M.R. and T.G. contributed to the draft of the article.

## Acknowledgment

We would like to thank the patients that participated in this study. We would like to thank the Cedars-Sinai Neurophysiology team that helped in EEG data acquisition and consent especially Cody Holland, Erica Quan and Ho Duong. We thank Karunesh Ganguly and Jaekyung Kim for NREM sleep oscillations detection Matlab code. The study was supported by American Heart Association (AHA; predoctoral fellowship 23PRE1018175 to R.R., postdoctoral fellowship 897265 to A.A. and career development award 847486 to T.G.), National Institutes of Health (NIH)’s National Institute for Neurological Disorders and Stroke (NINDS) (R00NS097620 and R01NS128469 to T.G.), National Science Foundation (award 2048231 to T.G.) and Cedars-Sinai Medical Center. A.A. also received support through Cedars-Sinai’s Center for Neural Science and Medicine postdoctoral fellowship.

## Data Availability Statement

The data that support the findings of this study are available from the corresponding author upon reasonable request.

## References

1. Ganguly K, Poo MM. Activity-dependent neural plasticity from bench to bedside. Neuron 2013;80(3):729–41.

2. Ganguly K, Khanna P, Morecraft RJ, Lin DJ. Modulation of neural co-firing to enhance network transmission and improve motor function after stroke. Neuron 2022;110(15):2363– 2385.

3. Norrving B, Kissela B. The global burden of stroke and need for a continuum of care. Neurology 2013;80:S5–12.

4. Gulati T, Guo L, Ramanathan DS, et al. Neural reactivations during sleep determine network credit assignment. Nature Neuroscience 2017;20:1277–1284.

5. Ramanathan DS, Gulati T, Ganguly K. Sleep-Dependent Reactivation of Ensembles in Motor Cortex Promotes Skill Consolidation. PLoS Biol 2015;13:e1002263.

6. Kim J, Gulati T, Ganguly K. Competing Roles of Slow Oscillations and Delta Waves in Memory Consolidation versus Forgetting. Cell 2019;179(2):514–526 e13.

7. Gulati T, Ramanathan DS, Wong CC, Ganguly K. Reactivation of emergent task-related ensembles during slow-wave sleep after neuroprosthetic learning. Nat Neurosci 2014;17(8):1107–13.

8. Genzel L, Kroes MC, Dresler M, Battaglia FP. Light sleep versus slow wave sleep in memory consolidation: a question of global versus local processes? Trends Neurosci 2014;37:10–19.

9. Stickgold R. Sleep-dependent memory consolidation. Nature 2005;437:1272–1278.

10. Tononi G, Cirelli C. Sleep and the price of plasticity: from synaptic and cellular homeostasis to memory consolidation and integration. Neuron 2014;81:12–34.

11. de Vivo L, Bellesi M, Marshall W, et al. Ultrastructural evidence for synaptic scaling across the wake/sleep cycle. Science 2017;355:507–510.

12. Klinzing JG, Niethard N, Born J. Mechanisms of systems memory consolidation during sleep. Nat Neurosci 2019;22(10):1598–1610.

13. Ebajemito JK, Furlan L, Nissen C, Sterr A. Application of Transcranial Direct Current Stimulation in Neurorehabilitation: The Modulatory Effect of Sleep. Frontiers in neurology 2016;7:54.

14. Kim J, Guo L, Hishinuma A, et al. Recovery of consolidation after sleep following stroke— interaction of slow waves, spindles, and GABA. Cell Reports 2022;38(9):110426.

15. Backhaus W, Braass H, Gerloff C, Hummel FC. Can Daytime Napping Assist the Process of Skills Acquisition After Stroke? Front Neurol 2018;9:1002.

16. Baumann CR, Kilic E, Petit B, et al. Sleep EEG Changes After Middle Cerebral Artery Infarcts in Mice: Different Effects of Striatal and Cortical Lesions. Sleep 2006;29(10):1339–1344.

17. Duss SB, Seiler A, Schmidt MH, et al. The role of sleep in recovery following ischemic stroke: A review of human and animal data. Neurobiology of Sleep and Circadian Rhythms 2017;2:94–105.

18. Facchin L, Schöne C, Mensen A, et al. Slow Waves Promote Sleep-Dependent Plasticity and Functional Recovery after Stroke. J. Neurosci. 2020;40(45):8637–8651.

19. Gao B, Cam E, Jaeger H, et al. Sleep Disruption Aggravates Focal Cerebral Ischemia in the Rat. Sleep 2010;33(7):879–887.

20. Giubilei F, Iannilli M, Vitale A, et al. Sleep patterns in acute ischemic stroke. Acta Neurologica Scandinavica 1992;86(6):567–571.

21. Gottselig JM, Bassetti CL, Achermann P. Power and coherence of sleep spindle frequency activity following hemispheric stroke. Brain 2002;125(2):373–383.

22. Poryazova R, Huber R, Khatami R, et al. Topographic sleep EEG changes in the acute and chronic stage of hemispheric stroke. Journal of Sleep Research 2015;24(1):54–65.

23. Siengsukon CF, Boyd LA. Sleep to learn after stroke: Implicit and explicit off-line motor learning. Neuroscience Letters 2009;451(1):1–5.

24. Lemke SM, Ramanathan DS, Darevksy D, et al. Coupling between motor cortex and striatum increases during sleep over long-term skill learning. eLife 2021;10:e64303.

25. Born J, Rasch B, Gais S. Sleep to Remember. Neuroscientist 2006;12(5):410–424.

26. Rothschild G, Eban E, Frank LM. A cortical–hippocampal–cortical loop of information processing during memory consolidation. Nat Neurosci 2017;20(2):251–259.

27. Sirota A, Csicsvari J, Buhl D, Buzsáki G. Communication between neocortex and hippocampus during sleep in rodents. Proceedings of the National Academy of Sciences 2003;100(4):2065–2069.

28. Walker MP, Brakefield T, Morgan A, et al. Practice with sleep makes perfect: sleep-dependent motor skill learning. Neuron 2002;35:205–211.

29. Buzsáki G. Hippocampal sharp wave-ripple: A cognitive biomarker for episodic memory and planning. Hippocampus 2015;25(10):1073–1188.

30. Cairney SA, Guttesen A á V, El Marj N, Staresina BP. Memory Consolidation Is Linked to Spindle-Mediated Information Processing during Sleep. Current Biology 2018;28(6):948–954.e4.

31. Latchoumane C-FV, Ngo H-VV, Born J, Shin H-S. Thalamic Spindles Promote Memory Formation during Sleep through Triple Phase-Locking of Cortical, Thalamic, and Hippocampal Rhythms. Neuron 2017;95(2):424–435.e6.

32. Fernandez LMJ, Lüthi A. Sleep Spindles: Mechanisms and Functions. Physiological Reviews 2020;100(2):805–868.

33. Cox R, Fell J. Analyzing human sleep EEG: A methodological primer with code implementation. Sleep Medicine Reviews 2020;54:101353.

34. Cox R. analyzing human sleep EEG [Internet]. 2020;[cited 2023 Sep 1] Available from: https://zenodo.org/record/3929730

35. Silversmith DB, Lemke SM, Egert D, et al. The Degree of Nesting between Spindles and Slow Oscillations Modulates Neural Synchrony. J Neurosci 2020;40(24):4673–4684.

36. Martínez-Cagigal V. Topographic EEG/MEG plot [Internet]. 2023;Available from: https://www.mathworks.com/matlabcentral/fileexchange/72729-topographic-eeg-meg-plot

37. Aarts E, Verhage M, Veenvliet JV, et al. A solution to dependency: using multilevel analysis to accommodate nested data. Nat Neurosci 2014;17(4):491–6.

38. Sullivan GM, Feinn R. Using Effect Size—or Why the P Value Is Not Enough. J Grad Med Educ 2012;4(3):279–282.

39. Cassidy JM, Wodeyar A, Wu J, et al. Low-Frequency Oscillations Are a Biomarker of Injury and Recovery After Stroke. Stroke 2020;51(5):1442–1450.

40. Carmichael ST, Chesselet MF. Synchronous neuronal activity is a signal for axonal sprouting after cortical lesions in the adult. J Neurosci 2002;22:6062–6070.

41. Gulati T, Won SJ, Ramanathan DS, et al. Robust neuroprosthetic control from the stroke perilesional cortex. The Journal of Neuroscience 2015;35:8653–61.

42. Tu-Chan AP, Natraj N, Godlove J, et al. Effects of somatosensory electrical stimulation on motor function and cortical oscillations. J NeuroEngineering Rehabil 2017;14(1):113.

43. van Dellen E, Hillebrand A, Douw L, et al. Local polymorphic delta activity in cortical lesions causes global decreases in functional connectivity. NeuroImage 2013;83:524–532.

44. Topolnik L, Steriade M, Timofeev I. Partial Cortical Deafferentation Promotes Development of Paroxysmal Activity. Cerebral Cortex 2003;13(8):883–893.

45. Helfrich RF, Mander BA, Jagust WJ, et al. Old Brains Come Uncoupled in Sleep: Slow Wave-Spindle Synchrony, Brain Atrophy, and Forgetting. Neuron 2018;97(1):221–230.e4.

46. Maingret N, Girardeau G, Todorova R, et al. Hippocampo-cortical coupling mediates memory consolidation during sleep. Nat Neurosci 2016;19(7):959–964.

47. Antony JW, Piloto L, Wang M, et al. Sleep Spindle Refractoriness Segregates Periods of Memory Reactivation. Current Biology 2018;28(11):1736–1743.e4.

48. Staresina BP, Bergmann TO, Bonnefond M, et al. Hierarchical nesting of slow oscillations, spindles and ripples in the human hippocampus during sleep. Nat Neurosci 2015;18(11):1679–1686.

49. Bergmann TO, Born J. Phase-Amplitude Coupling: A General Mechanism for Memory Processing and Synaptic Plasticity? Neuron 2018;97(1):10–13.

50. Peyrache A, Khamassi M, Benchenane K, et al. Replay of rule-learning related neural patterns in the prefrontal cortex during sleep. Nat Neurosci 2009;12:919–926.

51. Bhattacharya S, Donoghue JA, Mahnke M, et al. Propofol Anesthesia Alters Cortical Traveling Waves. Journal of Cognitive Neuroscience 2022;34(7):1274–1286.

52. Soplata AE, Adam E, Brown EN, et al. Rapid thalamocortical network switching mediated by cortical synchronization underlies propofol-induced EEG signatures: a biophysical model [Internet]. 2023;2022.02.17.480766.[cited 2023 Apr 10] Available from: https://www.biorxiv.org/content/10.1101/2022.02.17.480766v2

53. Bauerschmidt A, Al-Bermani T, Ali S, et al. Modern Sedation and Analgesia Strategies in Neurocritical Care. Curr Neurol Neurosci Rep 2023;

54. Kang Y, Saito M, Toyoda H. Molecular and Regulatory Mechanisms of Desensitization and Resensitization of GABAA Receptors with a Special Reference to Propofol/Barbiturate. Int J Mol Sci 2020;21(2):563.

55. Ouyang W, Wang G, Hemmings HC. Isoflurane and propofol inhibit voltage-gated sodium channels in isolated rat neurohypophysial nerve terminals. Mol Pharmacol 2003;64(2):373– 381.

56. Tang P, Eckenhoff R. Recent progress on the molecular pharmacology of propofol. F1000Res 2018;7:123.

57. Kondili E, Alexopoulou C, Xirouchaki N, Georgopoulos D. Effects of propofol on sleep quality in mechanically ventilated critically ill patients: a physiological study. Intensive Care Med 2012;38(10):1640–1646.

58. Yue X-F, Wang A-Z, Hou Y-P, Fan K. Effects of propofol on sleep architecture and sleep– wake systems in rats. Behavioural Brain Research 2021;411:113380.

59. Lynch BA, Lambeng N, Nocka K, et al. The synaptic vesicle protein SV2A is the binding site for the antiepileptic drug levetiracetam. Proc Natl Acad Sci U S A 2004;101(26):9861–9866.

60. Luz Adriana PM, Blanca Alcira RM, Itzel Jatziri CG, et al. Effect of levetiracetam on extracellular amino acid levels in the dorsal hippocampus of rats with temporal lobe epilepsy. Epilepsy Research 2018;140:111–119.

61. Bell C, Vanderlinden H, Hiersemenzel R, et al. The effects of levetiracetam on objective and subjective sleep parameters in healthy volunteers and patients with partial epilepsy. Journal of Sleep Research 2002;11(3):255–263.

62. Bazil CW, Battista J, Basner RC. Effects of levetiracetam on sleep in normal volunteers. Epilepsy & Behavior 2005;7(3):539–542.

63. Murphy M, Bruno M-A, Riedner BA, et al. Propofol Anesthesia and Sleep: A High-Density EEG Study. Sleep 2011;34(3):283–291.

64. Yeh W-C, Lu S-R, Wu M-N, et al. The impact of antiseizure medications on polysomnographic parameters: a systematic review and meta-analysis. Sleep Medicine 2021;81:319–326.

65. Purdon PL, Pierce ET, Mukamel EA, et al. Electroencephalogram signatures of loss and recovery of consciousness from propofol. Proc Natl Acad Sci U S A 2013;110(12):E1142–1151.

66. Bernhardt J, Hayward KS, Kwakkel G, et al. Agreed Definitions and a Shared Vision for New Standards in Stroke Recovery Research: The Stroke Recovery and Rehabilitation Roundtable Taskforce. Neurorehabil Neural Repair 2017;31(9):793–799.

67. Pearson-Fuhrhop KM, Kleim JA, Cramer SC. Brain Plasticity and Genetic Factors. Topics in Stroke Rehabilitation 2009;16(4):282–299.

68. Ganguly K, Byl NN, Abrams GM. Neurorehabilitation: Motor recovery after stroke as an example. Annals of Neurology 2013;74(3):373–381.

69. Dromerick AW, Lang CE, Birkenmeier RL, et al. Very Early Constraint-Induced Movement during Stroke Rehabilitation (VECTORS): A single-center RCT. Neurology 2009;73(3):195–201.

70. Dromerick AW, Geed S, Barth J, et al. Critical Period After Stroke Study (CPASS): A phase II clinical trial testing an optimal time for motor recovery after stroke in humans. Proceedings of the National Academy of Sciences 2021;118(39):e2026676118.

71. Krakauer JW, Carmichael ST, Corbett D, Wittenberg GF. Getting Neurorehabilitation Right: What Can Be Learned From Animal Models? Neurorehabil Neural Repair 2012;26(8):923–931.

72. Clarkson AN, Huang BS, MacIsaac SE, et al. Reducing excessive GABA-mediated tonic inhibition promotes functional recovery after stroke. Nature 2010;468(7321):305–309.

73. He W-M, Ying-Fu L, Wang H, Peng Y-P. Delayed treatment of α5 GABAA receptor inverse agonist improves functional recovery by enhancing neurogenesis after cerebral ischemia-reperfusion injury in rat MCAO model. Sci Rep 2019;9(1):2287.

74. Marshall L, Helgadottir H, Molle M, Born J. Boosting slow oscillations during sleep potentiates memory. Nature 2006;444:610–613.

75. Ngo H-VV, Martinetz T, Born J, Mölle M. Auditory Closed-Loop Stimulation of the Sleep Slow Oscillation Enhances Memory. Neuron 2013;78(3):545–553.

76. Levy RM, Harvey RL, Kissela BM, et al. Epidural Electrical Stimulation for Stroke Rehabilitation: Results of the Prospective, Multicenter, Randomized, Single-Blinded Everest Trial. Neurorehabil Neural Repair 2016;30(2):107–119.

77. Khanna P, Totten D, Novik L, et al. Low-frequency stimulation enhances ensemble co-firing and dexterity after stroke [Internet]. Cell 2021;Available from: https://www.ncbi.nlm.nih.gov/pubmed/33571430

78. Ramanathan DS, Guo L, Gulati T, et al. Low-frequency cortical activity is a neuromodulatory target that tracks recovery after stroke. Nat Med 2018;24(8):1257–1267.

79. Abbasi A, Danielsen NP, Leung J, et al. Epidural cerebellar stimulation drives widespread neural synchrony in the intact and stroke perilesional cortex. Journal of NeuroEngineering and Rehabilitation 2021;18(1):89.

80. Guo L, Kondapavulur S, Lemke SM, et al. Coordinated increase of reliable cortical and striatal ensemble activations during recovery after stroke. Cell Reports 2021;36(2):109370.

