## Supplementary Information for "Disturbed laterality of non-rapid eye movement sleep oscillations in post-stroke human sleep: a pilot study"

The supplementary information below includes a table on the statistical details of stroke versus contralateral mirrored (CM) electrodes' NREM oscillations comparisons (**Supp. Table 1**), and stroke versus contralateral non-mirrored (CNM) electrodes' NREM oscillations comparisons (**Supp. Table 2**); and a table on one-way ANOVA results for just stroke electrodes comparison in 3 medication-based groupings (**Supp. Table 3**). Supplementary figure (**Supp. Fig. 1**) shows the topographical density plots for different NREM oscillations with each panel with specific colormap scale for easier visualization of trends. Supplementary figure (**Supp. Fig. 2**) is included at the end that compares the NREM oscillations' densities for different patient groups on stroke verses contralateral non-mirror (CNM) electrodes.

| NREM oscillation density | Fixed-effects coefficients (95% CIs) | | | | Random effects covariance parameters (95% CIs) | | | | | | Model | |
|---|---|---|---|---|---|---|---|---|---|---|---|---|
| | Intercept | | Electrode | | Intercept | Electrode | Medication group | Intercept−Electrode | Intercept−Medication group | Electrode−Medication group | | |
| | ***$tStat_{42}$*** | ***p value*** | ***$tStat_{42}$*** | ***p value*** | ***std*** | ***std*** | ***std*** | ***corr.*** | ***corr.*** | ***corr.*** | ***Cohen's d*** | ***$R^2$*** |
| **Spindle** | 6.9079 | 1.9688 x10$^{-8}$ | 0.85155 | 0.39929 | 1.7457 | 1.45640 | 0.54078 | –0.95336 | –0.61764 | 0.82622 | 0.5651 | 0.2719 |
| **SO** | 7.2316 | 6.7928 x10$^{-9}$ | 3.0559 | 0.00389 | 0.6636 | 0.50056 | 0.46378 | –1 | –1 | 1 | 0.5346 | 0.2582 |
| **Delta (δ)** | 5.4601 | 2.3645 x10$^{-6}$ | 3.6979 | 0.00063 | 5.0144 | 2.90430 | 2.559 | –1 | –1 | 1 | 0.7788 | 0.3629 |
| **Nested SO-Spindle** | 7.1156 | 9.939 x10$^{-9}$ | 0.82454 | 0.41429 | 0.1458 | 0.18701 | 0.14452 | –0.99279 | –0.2272 | 0.34227 | 0.6823 | 0.3229 |
| **Nested δ-Spindle** | 5.6176 | 1.4069 x10$^{-6}$ | 0.56857 | 0.57268 | 1.0624 | 0.98551 | 0.43146 | –0.9972 | –0.56061 | 0.621 | 0.9031 | 0.4115 |

**Supplementary Table 1.** Linear mixed effect model results for stroke vs contralateral mirrored (CM) electrode analysis. $tStat_{df}$: t-statistic and df: degree of freedom; std: standard deviation; corr.: correlation; $R^2$: coefficient of determination.

| NREM oscillation density | Fixed-effects coefficients (95% CIs) | | | | Random effects covariance parameters (95% CIs) | | | | | | Model | |
|---|---|---|---|---|---|---|---|---|---|---|---|---|
| | Intercept | | Electrode | | Intercept | Electrode | Medication group | Intercept−Electrode | Intercept−Medication group | Electrode−Medication group | | |
| | ***$tStat_{38}$*** | ***p value*** | ***$tStat_{38}$*** | ***p value*** | ***std*** | ***std*** | ***std*** | ***corr.*** | ***corr.*** | ***corr.*** | ***Cohen's d*** | ***$R^2$*** |
| **Spindle** | 6.3677 | $1.7844 \times 10^{-7}$ | 1.9379 | 0.060086 | 3.1972 | 2.6202 | 1.4867 | -0.96893 | -0.91878 | 0.98787 | 0.5043 | 0.2445 |
| **SO** | 5.5363 | $2.4625 \times 10^{-6}$ | 3.5961 | 0.00091675 | 1.3433 | 0.99767 | 0.85578 | –1 | –1 | NaN | 0.5522 | 0.2662 |
| **Delta (δ)** | 4.9445 | $1.579 \times 10^{-5}$ | 3.9165 | 0.00036151 | 6.905 | 4.4912 | 4.0991 | –1 | –1 | 1 | 0.6900 | 0.3261 |
| **Nested SO-Spindle** | 6.1161 | $3.9458 \times 10^{-7}$ | 1.8632 | 0.070179 | 0.42091 | 0.41908 | 0.23671 | -0.99061 | -0.95263 | 0.98526 | 0.6246 | 0.2981 |
| **Nested δ-Spindle** | 6.1211 | $3.8835 \times 10^{-7}$ | 1.8995 | 0.065103 | 1.8198 | 1.6238 | 0.93552 | -0.99626 | -0.95183 | 0.97478 | 0.6374 | 0.3036 |

**Supplementary Table 2.** Linear mixed effect model results for stroke vs contralateral non-mirrored (CNM) electrode analysis. $tStat_{df}$: t-statistic and df: degree of freedom; std: standard deviation; corr.: correlation; $R^2$: coefficient of determination.

| NREM oscillation density | Group | | | Error | | | *F* | *p* |
|---|---|---|---|---|---|---|---|---|
| | SS | df | MS | SS | df | MS | | |
| **Spindle** | 44.844 | 2 | 22.4218 | 137.218 | 19 | 7.22 | 3.1 | 0.0681 |
| **SO** | 7.8303 | 2 | 3.91514 | 82.5297 | 19 | 4.34367 | 0.9 | 0.4227 |
| **Delta (δ)** | 106.01 | 2 | 53.0062 | 1041.95 | 19 | 54.8394 | 0.97 | 0.3983 |
| **Nested SO-Spindle** | 0.53641 | 2 | 0.26821 | 4.3391 | 19 | 0.23153 | 1.16 | 0.3352 |
| **Nested δ-Spindle** | 5.7491 | 2 | 2.87454 | 37.112 | 19 | 1.95326 | 1.47 | 0.2546 |

**Supplementary Table 3.** One-way ANOVA results for stroke electrode analysis. SS: sum of squares; df: degree of freedom; MS: mean square; *F*: *F*-statistic (ratio of two MS); *p*: significance values.

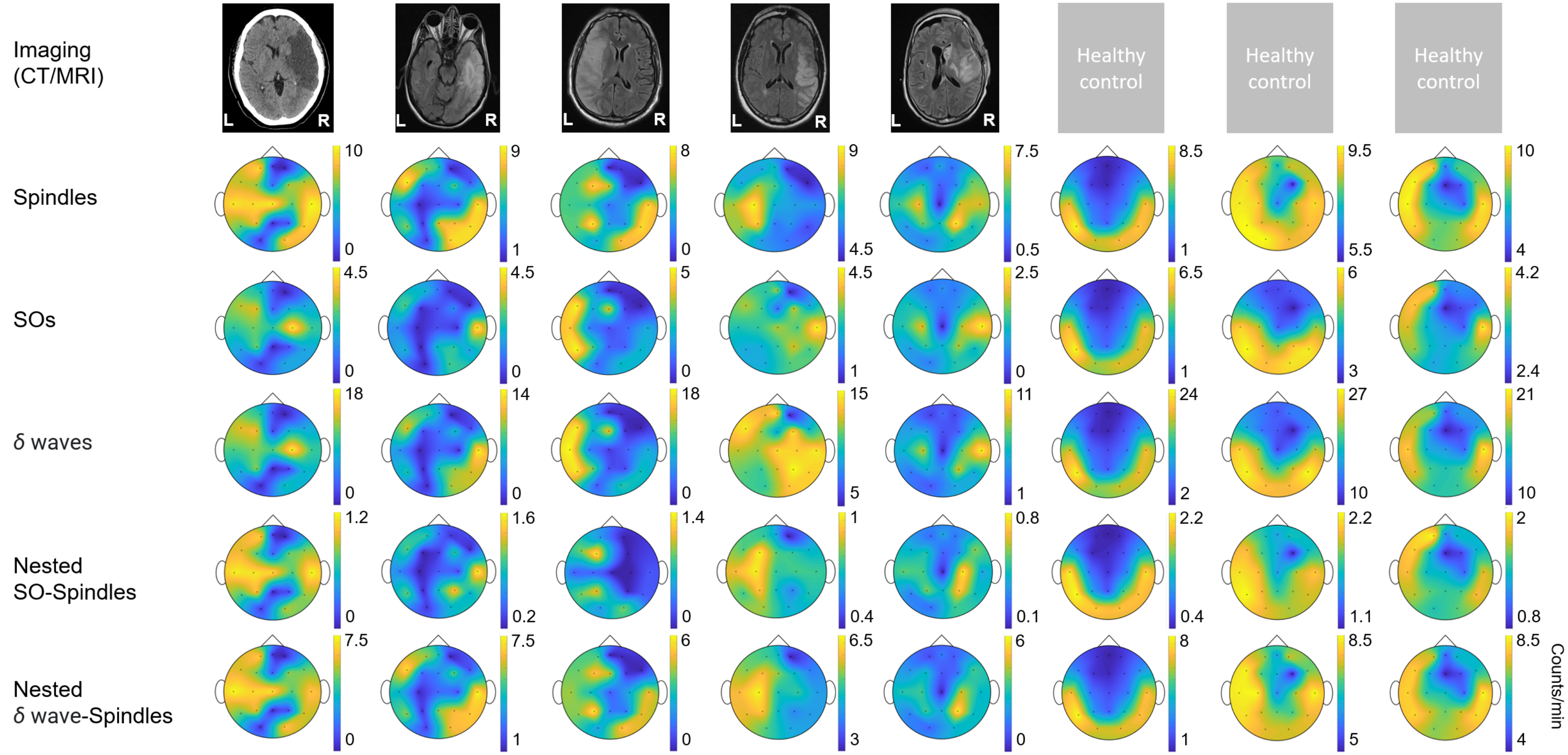


**Supplementary Figure 1. Imaging data and topographical density plots for different NREM oscillations.** Top to bottom: ***Imaging data***: CT (computed tomography) image for patient P1, T2 sequences of MRI (magnetic resonance imaging) images for patients P2 to P5; no imaging data available for healthy subjects (P6 to P8). Radiologic imaging has been flipped horizontally to align with topographic density maps, *i.e.*, image left, and right are ipsilateral to patient left and right. Left and right are marked in imaging figures (P1-P5) and apply to density topographical maps below them; ***Topographical maps*** for detected ***spindle*** density (count/min) during NREM sleep for all subjects; ***Topographical maps*** for detected ***SO*** density (count/min) during NREM sleep for all subjects; ***Topographical maps*** for detected ***δ waves'*** density (count/min) during NREM sleep for all subjects; ***Topographical maps*** for detected ***nested SO-spindle*** density (count/min) during NREM sleep for all subjects; ***Topographical maps*** for detected ***δ wave-nested-spindle*** density (count/min) during NREM sleep for all subjects. Colormap scale shown at right individually for each topographical plot.

### Stroke vs contralateral non-mirror (CNM) electrode comparison

**A**

| Patient | P1 | P2 | P3 | P4 | P5 |
|---|---|---|---|---|---|
| **Stroke electrodes** | C4,F8, T4,T6/ P8 | T4,P4, T6/P8 | C3,F7, P3,T3, O1,T5 /P7,F3 | P4,F8, T4,T6/ P8 | C4,F8, T4,T6/ P8 |
| **Contralateral non-mirror (CNM) electrodes** | F3,P3, O1,Fp 1 | F3,C3, O1,Fp 1,F7 | Fp2 | F3,C3, O1,Fp 1 | F3,P3, O1,Fp 1 |

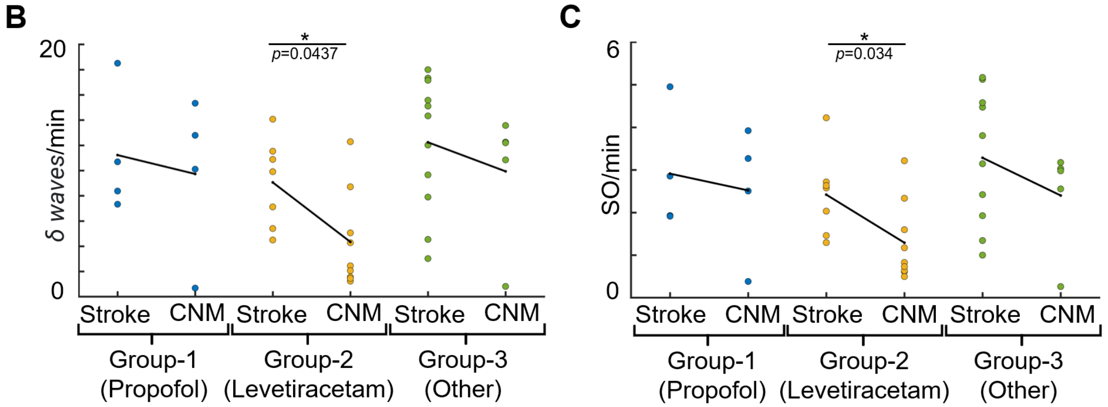


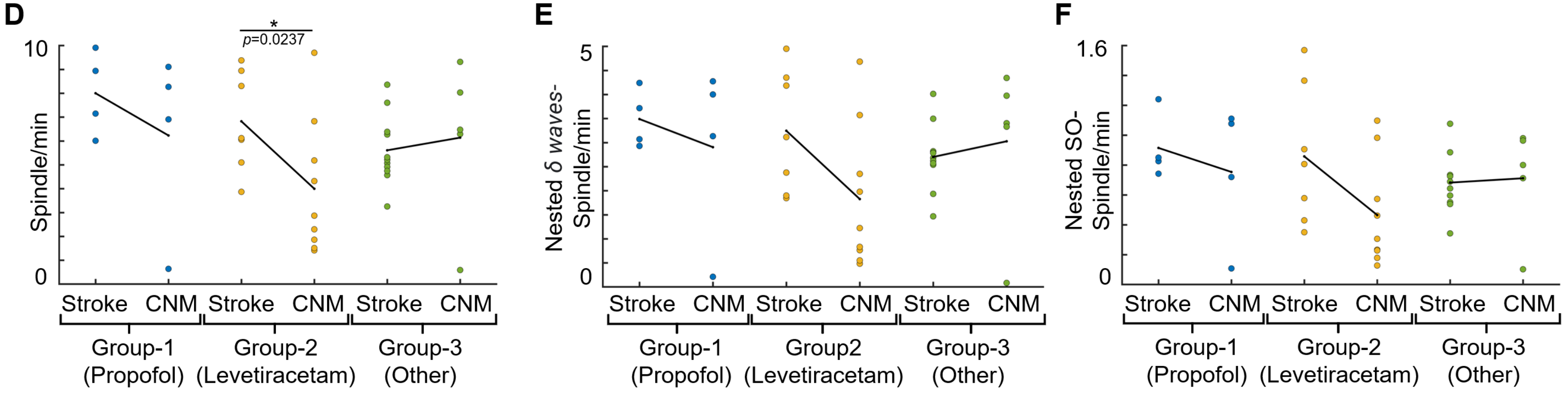


**Supplementary Figure 2. NREM oscillations' densities for different patient groups on stroke verses contralateral non-mirror (CNM) electrodes.** ***A*,** Table showing selected *stroke* and *contralateral non-mirror electrodes* (CNM) for all patients. ***B*,** Comparison of *δ wave* density (count/min) on *stroke versus CNM electrode*s for patients on different medications. Black line shows the mean values within the group. Dots represent the NREM oscillations' density for single electrode. ***C*,** Same as ***B*** for SO density. ***D*,** Same as ***B*** for spindle density. ***E*,** Same as ***B*** for nested *δ* wave-nested spindle density. ***F*,** Same as ***B*** for SO-nested spindle density. *: statistically significant *p* values for two-tailed *t*-test.
